# Genomic Epidemiology of *Enterococcus faecium* Bloodstream Infections During a VanB-type VRE Peak Reveals an Oligoclonal Scenario: An Observational Study at a German University Hospital (2017–2022)

**DOI:** 10.1101/2025.07.09.25331181

**Authors:** Leonard Knegendorf, Anna Sommer, Claas Baier, Robert E. Weber, Martin A. Fischer, Guido Werner, Stefan Ziesing, Dirk Schlüter

**Affiliations:** Hannover Medical School (MHH), Institute for Medical Microbiology and Hospital Epidemiology, Hannover, Germany; TWINCORE Centre for Experimental and Clinical Infection Research, Institute for Molecular Bacteriology, Hannover, Germany; Robert Koch Institute, Department of Infectious Diseases, Wernigerode Branch, Wernigerode, Germany; National Reference Centre for Staphylococci and Enterococci, Wernigerode, Germany; Infection Prevention and Control Department, Agaplesion Diakonieklinikum Rotenburg, Rotenburg (Wümme), Germany; Cluster of Excellence RESIST (EXC 2155), Hannover Medical School, Hannover, Germany

**Keywords:** Enterococcus faecium, vancomycin resistance, bloodstream infection, genomic epidemiology, split k-mer analysis

## Abstract

**Background:** A substantial and rapid increase, followed by a sharp decline in *vanB*-type vancomycin-resistant *E. faecium* (VRE), occurred in Germany in the late 2010s. This unusual epidemiological trend prompted detailed genomic investigations to explore the underlying dynamics at a German university hospital.

**Methods:** We retrospectively analyzed 344 *E. faecium* bloodstream infection (BSI) isolates collected between 2017 and 2022. Isolates were classified as *vanA*-positive, *vanB*-positive, or *van-*negative, while VRE was defined as *vanA*– or *vanB*-positive. Molecular typing included multilocus sequence typing (MLST), core genome MLST (cgMLST), and split k-mer analysis (SKA). Clonal relationships and potential patient-to-patient transmission events were assessed by integrating genomic data with machine-aided analysis of available longitudinal patient movement data.

**Results:** High-resolution genomic analysis of BSI isolates revealed an oligoclonal scenario involving multiple epidemic lineages (e.g., sequence type (ST)117/complex type (CT)71, ST117/CT929, ST117/CT2505, ST80, and ST262) with distinct *van* genotypes and dynamic changes over time. Overall, genomic overlap between *vanA*-positive, *vanB*-positive, and *van*-negative populations was minimal. The *vanB* surge, peaking in late 2018-early 2019, was mainly driven by ST117/CT71, ST117/CT36, and ST117/CT1917. SKA provided enhanced discriminatory power over cgMLST. The inferred transmission events linked bloodstream infections separated by long intervals (median, 106 days).

**Conclusions:** Accurate characterization of the transient *vanB*-type VRE peak required an integrated genomic approach combining cgMLST, high-resolution SKA, and epidemiological data. These comprehensive methods enable infection prevention and control (IPC) teams to distinguish true outbreaks from apparent epidemiologic clusters of polyclonal isolates. This allows for accurate interpretation and precisely targeted interventions where evidence supports transmission.

**Importance:** Vancomycin-resistant *Enterococcus faecium* is an increasingly important nosocomial pathogen worldwide. Understanding its epidemiology and transmission dynamics is critical to effectively control its spread. This study documents shifting *E. faecium* populations within a high-risk hospital environment, including both vancomycin-resistant and vancomycin-susceptible bloodstream isolates. Virulence gene profiling demonstrated that hospital-associated variants predominated across all major lineages, independent of vancomycin resistance, indicating that hospital adaptation is a common feature of both resistant and susceptible populations. Detailed genomic analyses, combining core genome multilocus sequence typing with high-resolution split k-mer analysis, integrated with comprehensive epidemiological tracking, were critical to accurately depict complex epidemiological dynamics. This combined approach allows precise differentiation between monoclonal outbreaks and oligoclonal transmission, enabling more targeted infection prevention and control strategies.

## Introduction

Enterococci are commensals of the human intestinal microbiome. Under certain conditions, however, they can also cause infections, particularly in immunocompromised and critically ill patients. They are a significant cause of nosocomial bloodstream infections. Of the many known *Enterococcus* species, only two are of major medical significance: *Enterococcus faecium* and *E. faecalis*. In particular, *E. faecium* isolates are characterized by high environmental resistance, an enhanced tolerance to disinfectants, and a wide range of acquired antibiotic resistances (1, 2). Hospital-associated isolates of this species differ from those colonizing the intestines of humans and animals. Genome comparisons show that infectious isolates fall into three distinct genomic clades called A1, A2, and B (3), whereas human commensal isolates of clade B are suggested to be reassigned to another species called *E. lactis* (4, 5).

Vancomycin is a first-line agent for the treatment of infections caused by enterococci, particularly *E. faecium*. Acquired resistance to vancomycin primarily occurs in members of the species *E. faecium* and to a much lesser extent in *E. faecalis*. Of the eight different variants of acquired vancomycin resistance, the *vanA* and *vanB* variants are of major medical importance. The *vanA* gene cluster is predominantly localized on plasmids that can be transferred between different strains. In contrast, the *vanB* gene cluster is mainly part of the bacterial chromosome, but can also be conjugated between different strains (6, 7). Both types of acquired vancomycin resistance could be transferred by clonal expansion or horizontal transfer.

Both national and international surveillance programs and schemes report comparably high rates of VRE in Germany. While VRE-BSI showed a rapid increase up until 2020, more recent data indicate a subsequent decline in case numbers (8). Data from the National Reference Centre for Enterococci, along with various publications on patient-associated enterococci, confirm the predominant prevalence of *vanB*-type VRE in Germany during the increase in VRE-BSI up to 2020.

The incidence of VRE BSI in Germany is relatively low, but it doubled from 1.7 to a peak of 3.0 per 100,000 in 2021 before declining back to 1.7 in 2023 (8). It ranges from 0.1% to 10.6% in high-risk hospitalized patients (9). Whole-genome sequencing (WGS) enables detailed strain classification, with MLST assigning a sequence type (ST) from allelic profiles of seven housekeeping genes and cgMLST assigning a complex type (CT) from the unique combination of alleles across 1,423 conserved loci (10).

Genomic analyses of local and regional invasive VRE strains in Germany from 2010 to 2020 revealed the preferential spread of certain strain variants associated with *vanB*-type resistance. For instance, *E. faecium* isolates of ST117/CT71/*vanB* were found to be widespread throughout Germany, whereas isolates of ST80/CT1065/*vanB* predominantly spread regionally focusing on southern and western Germany (11). Local outbreaks may be dominated by unique strain types such as ST80/CT1013/*vanB* as recently described (12).

The objective of this study was to investigate the clonal structure and temporal dynamics of bloodstream-associated *E. faecium* during a transient VanB-type VRE peak. To this end, we retrospectively analyzed isolates from BSIs in patients treated at Hannover Medical School, a 1,500-bed tertiary university hospital in northern Germany that specializes, among other areas, in solid-organ transplantation. The isolates were collected from 2017 to 2022 to cover the observed increase and decrease of VRE prevalence in Germany as well as the impact of the SARS-CoV-2 pandemic. Isolates underwent WGS and molecular typing, followed by epidemiological tracking of phylogenetically related strains to gain a deeper understanding of *E. faecium* epidemiology.

## Materials and Methods

### Clinical Case Definition

From October 11, 2017, to February 10, 2022, we included every *E. faecium* isolate from blood cultures for which antimicrobial susceptibility testing was performed in routine diagnostics. We included at least one strain per patient as well as strains with distinct colony morphology or isolated more than seven days after the initial isolate. We performed retrospective identification of isolates by querying the laboratory information system database (M/Lab, DORNER Health IT Solutions, Müllheim, Germany) and manually reviewing available isolates in our strain collections, which the laboratory routinely maintains. The isolates originated from various hospital wards, including the pediatric clinics, thus ensuring a comprehensive representation across different clinical settings. Throughout the study period, all patients who tested positive for VRE were isolated in single rooms or cohorted, and staff wore personal protective equipment (gowns and gloves).

Ethics committee of Hannover Medical School gave ethical approval for this work (No. 10388_BO_K_2022). Since this was a retrospective quality-assuring study, Ethics Committee and Data Protection Commissioner of Hannover Medical School waived the need for informed consent.

### Bacterial Isolates and Susceptibility Testing

Blood samples were inoculated in BD BACTEC™ Plus Aerobic and BD BACTEC™ Lytic Anaerobic culture vials (BD, Heidelberg, Germany) and incubated in a BD BACTEC™ FX system according to the manufacturer’s instructions. When flagged positive, blood culture samples were subcultured on solid media following internal laboratory protocols. Species identification was performed using matrix-assisted laser desorption/ionization time-of-flight mass spectrometry (MALDI-TOF MS) with a Vitek MS system (bioMérieux, Marcy-l’Étoile, France). Antimicrobial susceptibility testing (AST) was conducted with a Vitek 2 system (bioMérieux, Marcy-l’Étoile, France). A VRE was identified when the isolate tested resistant to vancomycin according to the European Committee on Antimicrobial Susceptibility Testing (EUCAST) breakpoints that were valid for the respective year of the study. Furthermore, a VanA phenotype was characterized by resistance to vancomycin and teicoplanin, while a VanB phenotype was characterized by resistance to vancomycin, and susceptibility to teicoplanin. Bacterial strains were stored in tryptone soy broth (Oxoid, Wesel, Germany) supplemented with 30% glycerine (CHEMSOLUTE©, Th. Geyer, Renningen, Germany) and kept at -80°C until whole-genome sequencing.

### Whole-genome Sequencing, Quality Control and Genome Reconstruction

Bacterial cultures were grown in Brain Heart Infusion (BHI) broth. Genomic DNA was extracted from an overnight culture using the DNeasy Blood & Tissue Kit (Qiagen, Hilden, Germany). DNA concentration was quantified with the Qubit dsDNA HS Assay Kit on a Qubit 4 Fluorometer (Thermo Fisher Scientific, Karlsruhe, Germany). Sequencing libraries were prepared using the Nextera XT DNA Library Preparation Kit (Illumina, San Diego, CA, United States), and paired-end sequencing was conducted on a NextSeq instrument with a read length of 150 bp (Illumina, San Diego, CA, United States). Raw sequence data quality was assessed using FQStat v0.0.12 (13), FastQC v0.12.1 (Andrews *et al*., https://www.bioinformatics.babraham.ac.uk/projects/fastqc/), AQUAMIS v1.3.12 (14) and SeqSphere^+^ v9.0.10 (Ridom, Münster, Germany). Eleven of 344 isolates were excluded due to inadequate quality control parameters and were not included in further analyses. These included six with AQUAMIS failure, one with <95% good cgMLST targets, and four due to multiple reasons including AQUAMIS failure. After quality control, Illumina raw reads were *de novo* assembled using SPAdes v3.14.1 with default parameters (15).

### Resistance and Virulence Gene Identification

The identification of the glycopeptide resistance genes *vanA* and *vanB* was performed using *de novo* assembled contigs with CLC Genomics Workbench v24.0.1 and the ‘Find Resistance with Nucleotide Database’ tool utilizing the QMI-AR Nucleotide Database.

Virulence gene screening was performed using VirulenceFinder for *Enterococcus faecium* and *Enterococcus lactis* (16) with BLAST-based identification at ≥90 % sequence identity and ≥60 % gene coverage. Genes of interest included adhesion and biofilm-associated factors such as the cell wall-anchored adhesin *ecbA* (17), the biofilm formation-associated proline-rich surface protein *prpA* (18), pilus gene cluster 1 (PCG-1) (19), and the hyaluronidase *hylEfm* (20), as well as the Toll/interleukin-1 receptor domain-containing immunomodulatory genes *tirE1* and *tirE2* (21). In addition, the bacteriocin gene *T8* was detected using a BLAST-based approach with the reference sequence GenBank accession no. DQ402539.1 and identical threshold criteria. Visualization of gene presence and variant calls followed the same layout, and graphical approach as Rubin *et al.* (22).

### Molecular Typing

MLST and cgMLST were carried out using established typing schemes in Ridom SeqSphere+ v9.0.10 (10, 23). Based on cgMLST, a Minimum Spanning Tree (MST) was inferred using the option ‘pairwise ignoring missing values’. CgMLST clusters were then defined by a pairwise allelic difference threshold of ≤15 alleles. In addition to cgMLST, split k-mer analysis (SKA, v1.0) was performed (24). Briefly, Split kmer files (.skf) were generated directly from paired raw FASTQ reads using *ska fastq* with default filtering parameters and a split k-mer size of 15. All files were summarized using *ska summary* for quality control, assessing the number of split k-mers and GC content to confirm expected genome representation, sequencing depth, and data integrity. Individual split k-mer files were then merged into a combined dataset using *ska merge*. Pairwise genomic distances between isolates were then calculated using *ska distance*, and clusters were assigned based on a 7-SNP cutoff and a minimum split k-mer identity of 90%.

### Epidemiological Analysis

For in-depth epidemiological analysis, we manually extracted data on patient movements from the hospital information system. Only inpatient stays were considered, and the analysis was limited to the ward level. An epidemiological link was defined as two patients having overnight stays on the same ward within a rolling 30-day window, with both stays preceding detection of the secondary patient’s bloodstream isolate. Cases that did not meet this criterion were not considered epidemiologically linked. To systematically identify such links, a machine-aided approach was implemented using Python (v3.13.3) and pandas (v2.2.3). We programmatically queried patient movement data to identify qualifying patient-to-patient contacts based on the defined criteria and the sampling dates of the corresponding bloodstream isolates. Information about VRE colonization was retrieved manually from the laboratory information system.

To aid the interpretation of potential transmission routes, we stratified clinical departments into three specialty groups based on the organ systems predominantly treated: (i) hematopoietic system, including hematology-oncology and hematopoietic stem cell transplantation units; (ii) excretory system, encompassing gastroenterology, hepatology, nephrology, urology, visceral surgery, and related fields; and (iii) respiratory and cardiovascular system, including pneumology, cardiology, cardiac surgery, and associated specialties. These groupings were used to reflect likely overlap in patient pathways and potential transmission reservoirs across functionally related wards. Departments that did not clearly align with one of these three categories were assigned to an “Other” group. In cases where patients were treated in interdisciplinary intensive care units, stratification was based on the underlying disease rather than the unit type.

Statistical analysis was performed using GraphPad Prism v10.5.0 (GraphPad Software, Boston, MA, United States).

## Results

### The Rise and Fall of VanB type VRE at Hannover Medical School

The prevalence of VRE in patients at Hannover Medical School was assessed from 2013 to 2024_1 (suffix _1/_2 = first/second half-year). Following a sharp increase beginning in 2017 and peaking in 2019, the absolute number of patients with the VanB phenotype steadily declined, reaching pre-2017 levels by 2022. This downward trend began well before the overall decline in cases treated at the hospital, which was largely attributable to the COVID-19 pandemic. In contrast, the number of VRE patients with the VanA phenotype has remained largely constant throughout the period (Fig. 1).

**Figure 1:**
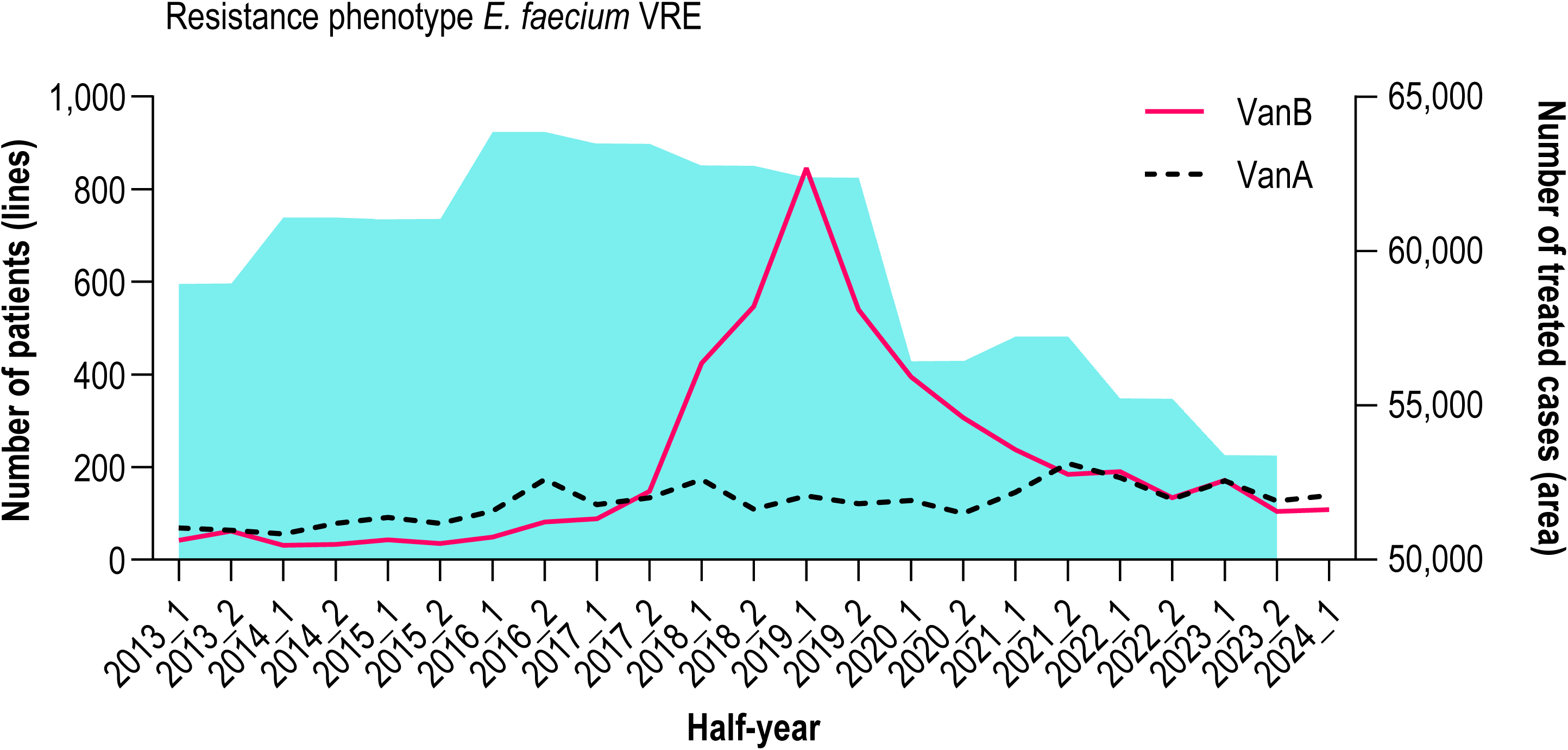
Phenotypic background of the VanB peak at Hannover Medical School from (2013 to 2024_1). The solid (VanB) and dashed (VanA) lines show the number of VRE patients per half-year from any specimen type (routine clinical or screening) across all hospital departments. The light-blue area indicates the total number of patients treated hospital-wide (cases) per half-year in the same interval.

### Isolate Collection Strategy

To analyze the temporal dynamics and genotype of *E. faecium* strains causing invasive disease, we only included *E. faecium* strains isolated from blood cultures between 2017 and 2022. Of 414 isolates (98 VRE, 316 vancomycin-susceptible (VSE)), 344 underwent whole-genome sequencing; 68 isolates were not stored due to process-related omissions in routine storage workflows. Following quality control, 333 isolates (80.43%) were included in the final analysis (Fig. 2A). The annual distribution of *E. faecium* isolates showed considerable variation, with the highest count recorded in 2021 (2017: n=5, 2018: n=61, 2019: n=52, 2020: n=92, 2021: n=102, and 2022: n=21; Fig. 2B). Among the isolates, 238 did not carry any *van* resistance genes, while 19 were positive for *vanA* and 76 for *vanB*. Due to the presence of the corresponding resistance determinants, the isolates were classified as VRE. For subsequent population analysis, duplicate isolates were excluded (same patient, same CT).

**Figure 2:**
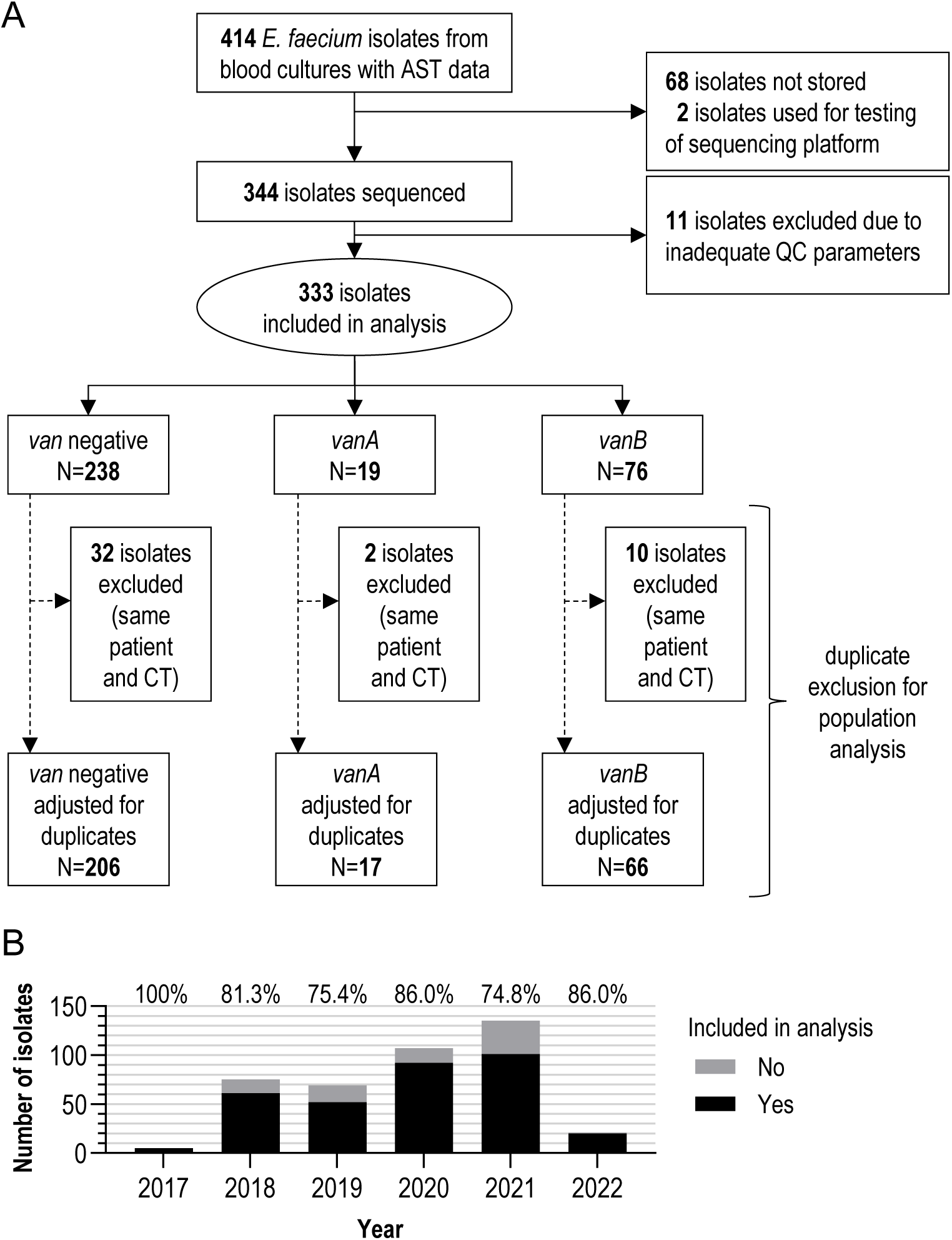
Overview of the isolates analyzed in this study. **(A)** Flowchart depicting the selection process of *E. faecium* isolates from BSIs. Exclusions due to unavailability, quality control issues, and duplicate cases are indicated, along with the stratification of isolates into *vanA*-, *vanB*-, and *van*-negative genotypes. **(B)** Proportion of isolates included in the final analysis relative to the total number of isolates meeting the inclusion criteria separated by year.

### Temporal shifts in clonal lineages and *van*-genotype prevalence

Population analysis based on MLST and cgMLST revealed temporal variations in the distribution of specific STs/CTs over time, with certain lineages, such as ST117/CT71/*vanB* and ST117/CT929/*van*-negative, persisting throughout the study period (Fig. 3A/B). From 2019 onward, however, ST117/CT71/*vanB* declined yet remained detectable, whereas ST117/CT929/*van*-negative and ST117/CT2505, *van*-negative and *vanA* variants, became increasingly prevalent in the following years.

**Figure 3:**
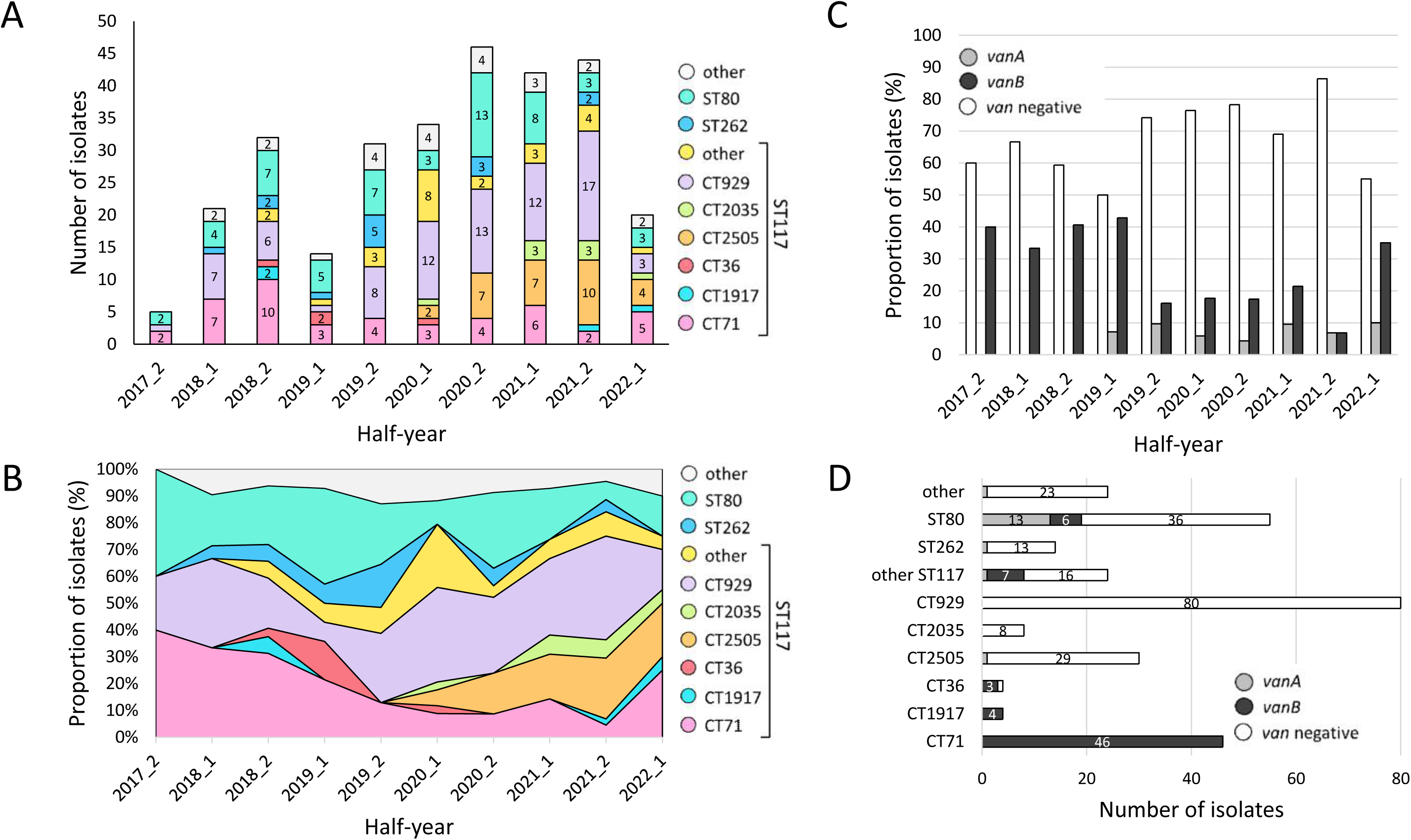
Characterization and temporal distribution of invasive VRE isolates at Hannover Medical School from 2017_2 to 2022_1. **(A)** Number of isolates per half-year, categorized by ST and CT. **(B)** Proportion of isolates assigned to each ST/CT over time. **(C)** Proportion of isolates with *vanA*, *vanB*, and *van*-negative genotypes over time. **(D)** Total number of isolates for each ST/CT, stratified by *vanA*, *vanB*, and *van*-negative genotypes.

The proportion of isolates with *vanA, vanB*, and *van-*negative genotypes varied throughout the study period. *Van*-negative isolates consistently accounted for the majority of analyzed isolates, with an increasing trend until 2021 (Fig. 3C). In this cohort, *vanA*-positive BSI isolates first appeared in 2019 and remained at low, fluctuating levels thereafter. After maintaining a stable prevalence of 30-40%, *vanB*-positive *E. faecium* BSI isolates declined sharply from the second half of 2019 (2019_2) onward to approximately 10% in subsequent years.

A detailed analysis of isolates for each *van-*genotype underscores the contribution of certain clonal lineages – such as ST117/CT71, ST117/CT36, and ST117/CT1917 – to the prevalence of *vanB-*positive isolates (Fig. 3D). These clonal types were particularly prevalent during the second half of 2018 (2018_2) and the first half of 2019 (2019_1; Fig. 1, Fig. 3B/3D). Among the *van*-negative isolates, ST117/CT929 was the most dominant, followed by ST80 (17 CTs, chiefly CT467 and CT1470) and ST117/CT2505. Notably, *vanA*-positive isolates were rare and primarily detected in ST80/CT1470.

### Prevalence of *van* genotypes and virulence genes across diverse cgMLST clusters

An MST was generated using cgMLST with a cluster threshold of ≤15 alleles difference. The total of 333 isolates was grouped into 17 distinct clusters, while 50 isolates remained as singletons (i.e., isolates not grouped into a cluster; Fig. 4A). Four out of the 17 clusters were designated major cgMLST clusters, which included the predominant clones ST117/CT929 and ST117/CT2505 (Cluster 1, n=111), ST117/CT71 (Cluster 2, n=47), ST80/CT467 (Cluster 3, n=23), and ST80/CT1470 (Cluster 4, n=15). In line with the analysis of *van* genotypes (Fig. 4D), Cluster 1 is primarily composed of *van*-negative *E. faecium* isolates, whereas Cluster 2 is largely associated with *vanB*-carrying variants (Fig. 4B). Although *vanA* was found in various clusters, Cluster 4 displayed the highest proportion of *vanA*-positive isolates.

**Figure 4:**
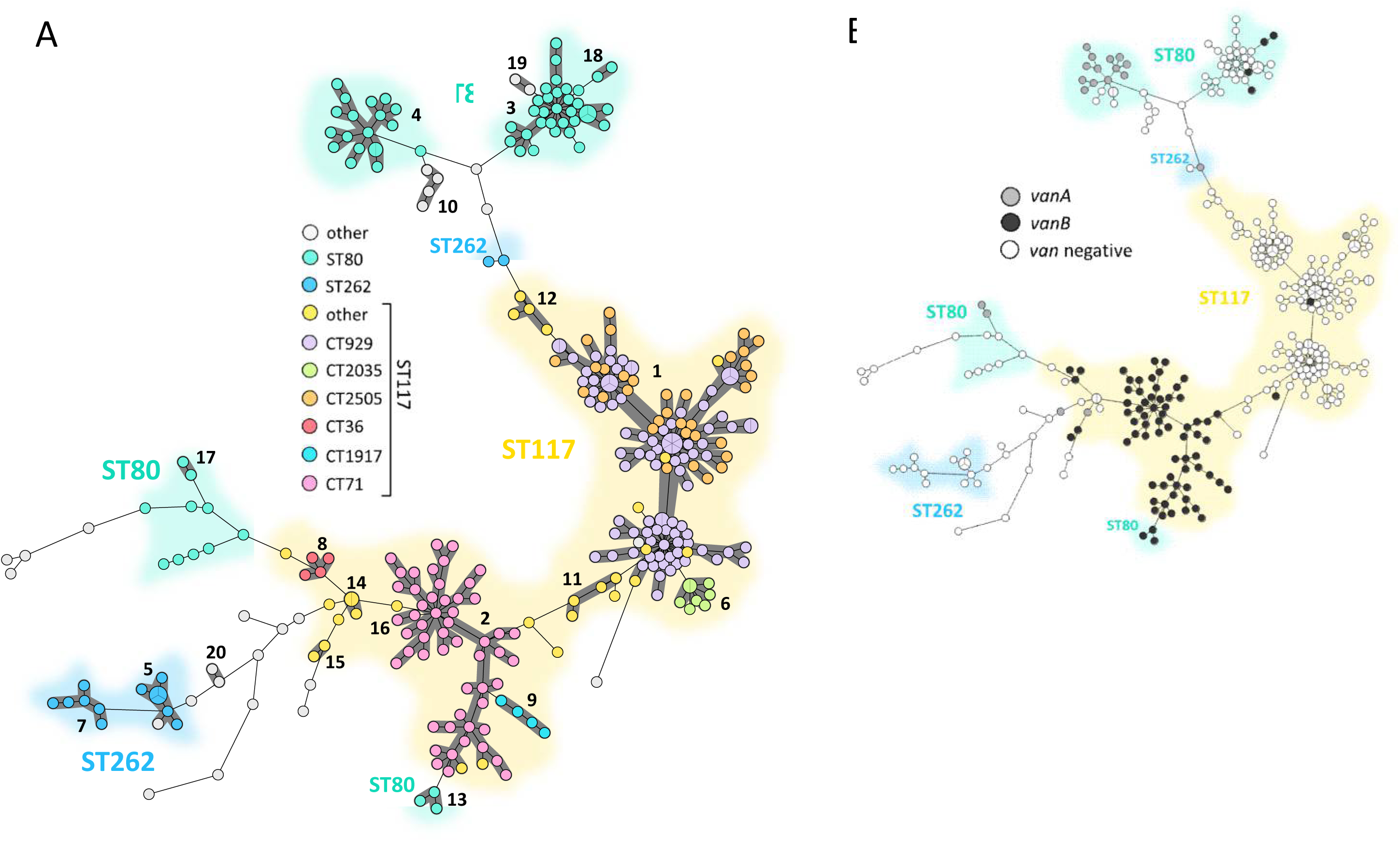
MST of invasive VRE isolates collected from 2017_2 to 2022_1 at Hannover Medical School. The MST was inferred based on cgMLST applying the “pairwise ignore missing values” option. **(A)** Distribution of STs and CTs. Clusters (shown as dark gray shaded areas and labeled with black cluster numbers) were defined using a pairwise allelic difference threshold of ≤15 alleles and are numbered by ascending size. **(B)** Distribution of *van* genotypes: *vanA* (gray), *vanB* (black), *van*-negative (white).

Virulence gene analysis revealed consistent detection of multiple virulence determinants across all major clonal lineages (Fig. S1). Where allelic differentiation was possible, hospital-associated (HA) variants predominated; only *prpA* showed a higher prevalence of community-associated (CA) alleles. *E. lactis*-like variants of the PGC-1 were exclusive to ST117/CT36, while PGC-1 was completely absent from ST117/CT71 and from ST117/CT1917 isolates collected in 2018. Separate virulence profiles were observed for ST117/CT1917 isolates emerging from 2021, corresponding to the two temporally distinct appearances of ST117/CT1917 during and after the *vanB* peak (Fig.□3). The genes *tirE1* and *tirE2* were detected in ST117/CT929, ST117/CT2505, ST117/CT71, ST117/CT2035, and in the isolates of ST117/CT1917 from 2021 onward. No isolate carried *hylEfm*. The bacteriocin gene *T8* was detected in 92.3% of isolates (Table S1).

### SKA provides higher resolution clustering than cgMLST

The assignment of individual isolates to specific clusters has important implications for the definition of outbreaks, hospital epidemiology, and the respective measures to prevent transmission of enterococci. Therefore, we tested different methods for defining clusters of *E. faecium* isolates, specifically focusing on cgMLST and SKA (Fig. 5). The classification of *E. faecium* isolates follows a stepwise refinement across the applied typing methods, progressively increasing resolution from MLST to cgMLST and SKA (Fig. 5). Starting with MLST, isolates were assigned to 16 STs, with ST117 (n=225), ST80 (n=66), and ST262 (n=16) being the most prevalent. While MLST provides only a broad genetic classification, cgMLST enhances resolution by reassigning the 16 identified STs into 70 CTs. Using a cluster threshold of ≤15 alleles, these 70 CT-defined groups were further consolidated into 20 possibly outbreak-related clusters and 44 singletons (see also Fig. 4). While cgMLST relies on 1,423 core genes, SKA, as a k-mer-based approach, also incorporates the accessory genome and intergenic regions. Due to this broader genomic inclusion, a considerable number of isolates classified within cgMLST outbreak clusters were reassigned as singletons in the SKA analysis. Using SKA, 40 clusters and 144 singletons were identified.

**Figure 5:**
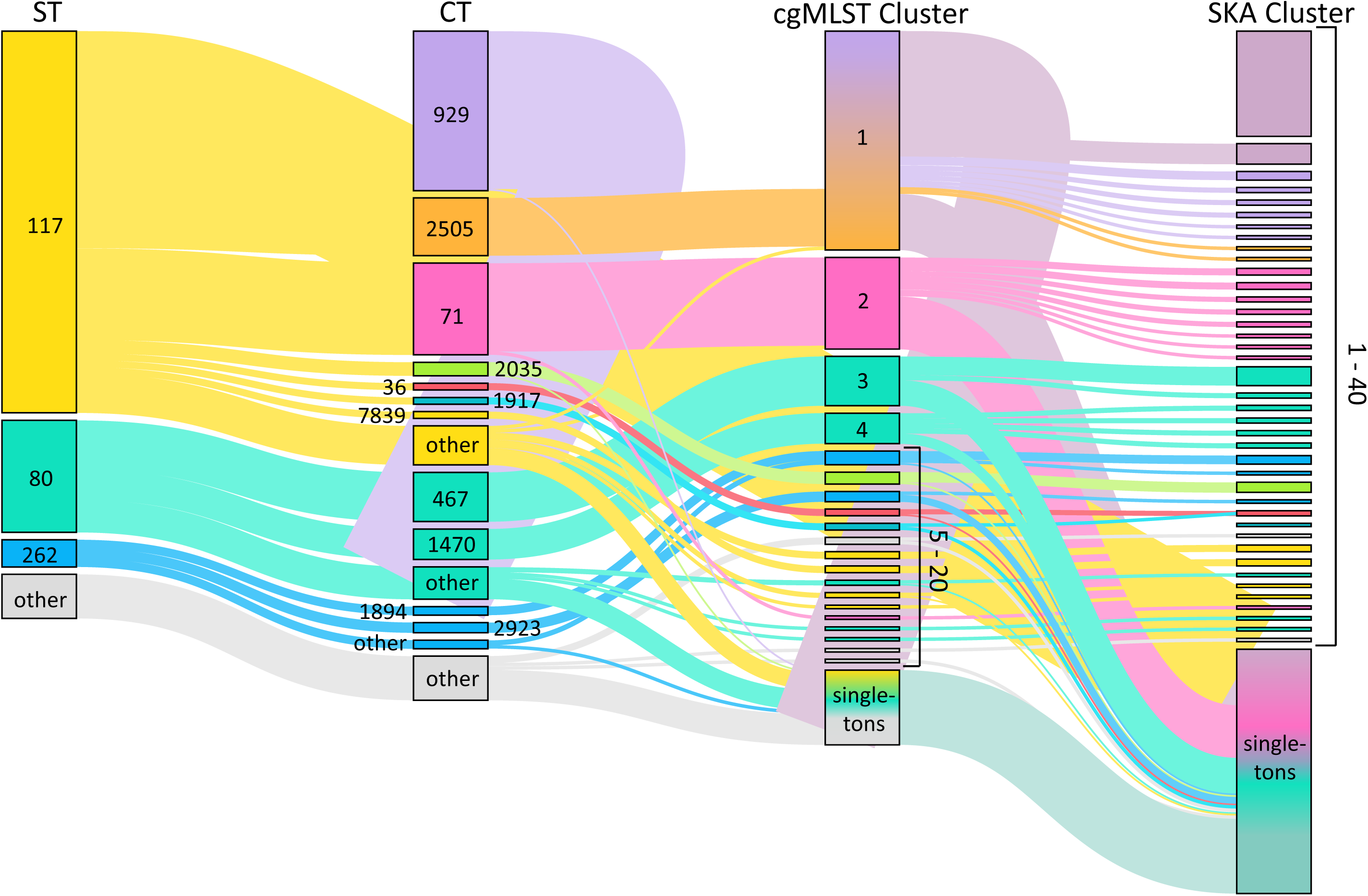
Hierarchical classification of *E. faecium* isolates based on different typing methods. CgMLST clusters were defined using a pairwise allelic difference threshold of ≤15 alleles. SKA clusters were defined by a 7-SNP threshold. Connecting lines indicate how individual isolates are assigned across typing methods. Node size corresponds to the number of isolates per category, and node labels indicate ST, CT, or cluster number, as designated by the column-specific headers above each group of nodes.

### Longitudinal Analysis of *E. faecium* ST117/CT71 Transmission

Understanding the epidemiological dynamics of *E. faecium* transmission is crucial for identifying potential outbreaks and improving infection control strategies. This analysis focuses on cgMLST cluster 2 (ST117/CT71) as one key driver of the *vanB* peak, examining spatial and temporal relationships to assess potential epidemiologic linkages.

A total of 52 ST117/CT71 isolates, one ST117/CT5101, and one ST117/CT7838 from 47 individual patients constituted the cgMLST cluster 2 (7 patients had two clinical isolates) (Fig. 6A). The majority of patients were treated in one of three organ system-based specialty groups (see Materials and Methods): hematopoietic system, excretory system, or cardiovascular and respiratory system (Fig. S2). Of the 47 patients, 24 (51.1%) had a documented intestinal VRE colonization prior to or at the time of infection. The cluster includes infection isolates from 2017-2022, with affected patients hospitalized across 36 wards.

**Figure 6:**
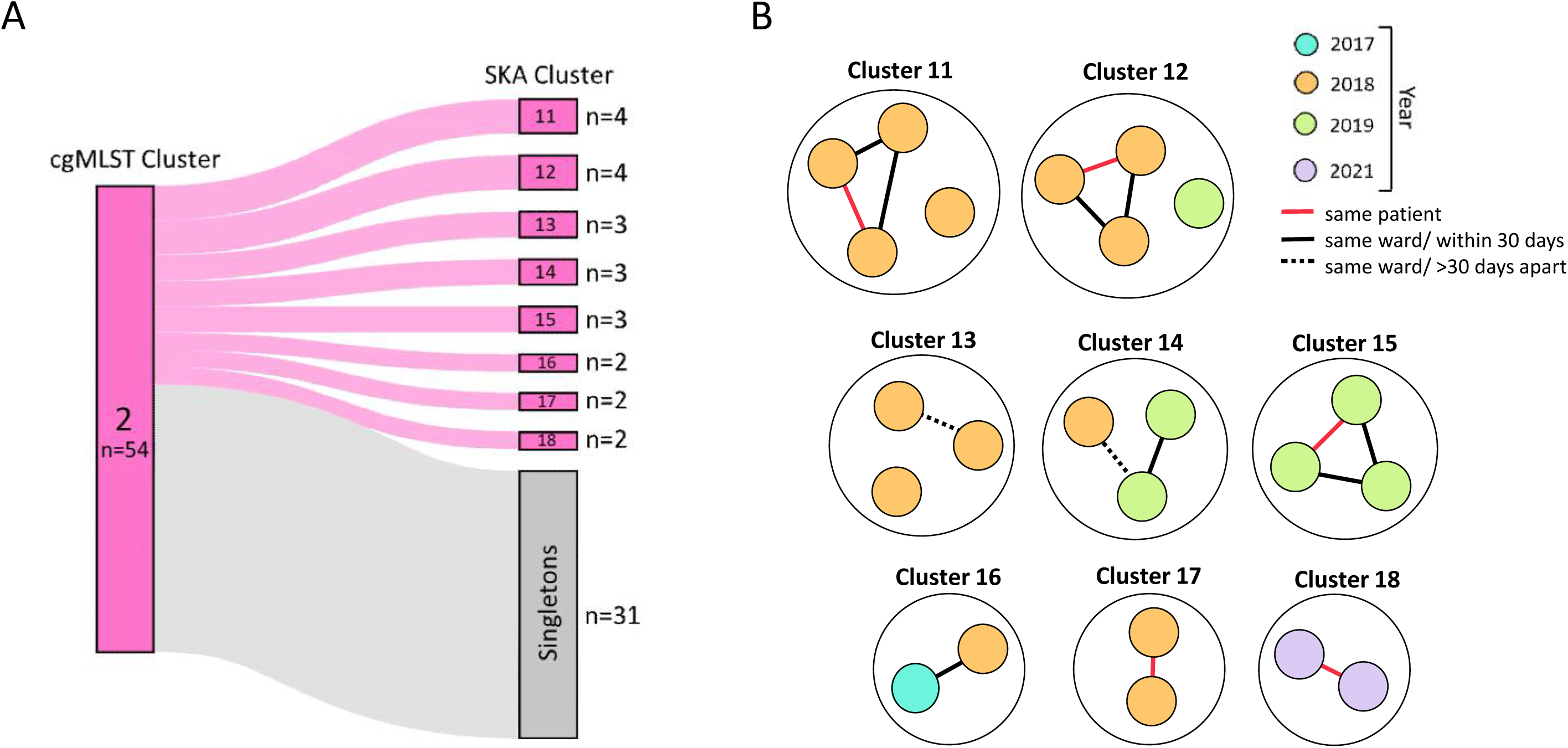
Identification of epidemiological links in SKA-defined outbreak clusters. **(A)** Sankey diagram illustrating how cgMLST cluster 2 partitions into distinct SKA clusters and singletons. **(B)** Visualization of epidemiological connections within SKA clusters. The colors of the circles represent the year of isolation. Lines indicate patient contact categories, based on defined ward-level overlaps.

SKA subdivided cgMLST cluster 2 into eight clusters and 31 singletons (Fig. 6A). Within all SKA clusters, epidemiological links could be detected, and five of the eight SKA clusters showed strong epidemiological linkage indicative of potential transmission (same ward within 30 days; cluster 11, 12, 14, 15, and 16; Fig. 6B). Five isolate pairs originated from the same patient (cluster 11, 12, 15, 17, and 18). We also analyzed available epidemiological data (Fig. S2) for potential links between all isolates in cgMLST cluster 2. Notably, 33 of 47 patients (70.2%) had been admitted to more than two different wards during the study. Our analysis identified 29 patients with epidemiological links within this cluster, despite their isolates being classified as singletons in SKA. Several patients were linked to others either as potential sources or recipients, with some having multiple plausible links, resulting in 55 total links across both SKA-defined clusters and singletons. The median time interval between BSIs across all links was 106 days (95% CI: 59.0-246.0). Notably, links in SKA-defined clusters showed a significantly shorter median interval between BSIs of 22 days compared to 118.5 days for links of SKA singletons (*p* = 0.0499, two-tailed Mann-Whitney *U* test).

## Discussion

The global emergence and dissemination of VRE remain pressing public health challenges, with notable epidemiological trends documented in Europe and worldwide. Germany experienced a marked increase in VRE cases during the late 2010s (25, 26), primarily driven by the clonal expansion of *vanB*-positive lineages, notably ST117/CT71 (27–29). Similar *vanA* to *vanB* dominance shifts have been observed globally, though circulating clones frequently remain region-specific (11, 30–32).

At Hannover Medical School, VanB*-*type VRE cases rose sharply between 2017 and 2019, peaked in 2019, then declined significantly and stabilized by 2021. For this study, we performed genomic population analysis on 80.43% of all *E. faecium* BSI isolates meeting the inclusion criteria during the study period, thus providing a robust representation of the clinical population. We identified an overall proportion of 23.67% vancomycin resistance, aligning with European reports (33, 34).

CgMLST analysis revealed that *vanB*-type rise and fall were predominantly associated with ST117/CT71 (31.3% in 2018_2; 21.4% in 2019_1) but also other clonal lineages like ST117/CT36 (14.3% in 2019_1) and ST117/CT1917 (6.3% in 2018_2). This aligns with findings from the National Reference Centre on Enterococci and others, which reported country-wide expansion of ST117/CT71/*vanB* until 2019, followed by significant decline (28, 29, 35). Similarly, Danish surveillance detected the emergence of ST117/CT36 starting in 2019, likely originating from German hospitals (32). Mills *et al.* demonstrated that emergent VRE lineages (ST80, ST117, ST1478) achieve competitive advantage through bacteriocin *T8* production, present in 70-96% of emergent lineages but only 16% of others, thereby providing significant colonization advantages *in vivo* (36). In our study, *T8* was detected in >90% of bloodstream isolates, confirming its widespread presence across dominant clonal lineages.

A tendency toward increased virulence gene content was observed in later-emerging lineages, such as ST117/CT2505 and ST117/CT2035. Comparison with Danish data published by Rubin *et al.* (22) supports this observation: whereas our ST117/CT36 isolates (mainly from 2018_2–2020) lacked *ecbA*, *hylEfm*, and PGC-1 and exhibited overall lower frequencies of adhesion– and colonization-associated genes, Danish ST117/CT36 isolates emerging from 2021 onward displayed a broader virulence gene repertoire. The immunomodulatory *tirE* genes, identified by Rubin *et al.* as specific for the latest emerging lineage ST80/CT2406 in Denmark, were consistently present in our latest emerging lineages ST117/CT2035 and ST117/CT1917 isolated from 2021 onward. The transient dominance of specific clones contrasts with Australian data showing stable virulome and resistome profiles of major clonal lineages over 15 years (37). In our study, even vancomycin-susceptible bloodstream isolates predominantly carried hospital-associated (HA) virulence alleles, indicating that key adaptation traits are not limited to VRE. Collectively, these findings suggest transmission efficiency drives lineage success rather than escalating virulence, consistent with the view that enterococcal virulence factors primarily serve ecological rather than pathogenic roles (38). While VRE now matches methicillin-resistant *Staphylococcus aureus* (MRSA) in disease burden, awareness remains disproportionately focused on MRSA (39).

The *vanB* peak did not coincide with an overall increase in BSIs, suggesting a shift in the underlying population structure – likely due to changes in colonizing strain prevalence – rather than an acute outbreak scenario. From mid-2019 onwards, *van*-negative isolates, primarily ST117/CT929 (33.7% in 2021) and ST117/CT2505 (19.8% in 2021), became increasingly prevalent. These lineages have been identified throughout Germany and rose notably from 2020 onward (35), with a recent study finding clonal isolates of these lineages being vancomycin-resistant (40). Although ST117/CT71/*vanB* isolates decreased significantly after 2019, they persisted throughout the study period. This suggests that there were ongoing transmission chains despite intensified infection control measures during the SARS-CoV-2 pandemic, which were expected to limit nosocomial transmission in general (41). These transmissions may have originated from environmental reservoirs, within or outside the hospital.

In contrast, *vanA*-type associated BSIs – absent during the peak of *vanB* – emerged from 2019 onward, mainly driven by *vanA*-type ST80 expansion. Recent reports demonstrate rapid national and cross-border dissemination of new *vanA*-positive lineages (32, 42, 43). Alongside the generally more stable prevalence of the VanA phenotype, these observations highlight the complexity of VRE resistance dynamics and underscore the importance of continued molecular surveillance.

Although horizontal transfer of vancomycin-resistance genes in *E. faecium* is possible (7), *van*-positive isolates rarely cluster with *van*-negative isolates, indicating limited transmission of corresponding resistance determinants between vancomycin-resistant and VSE BSI isolates, confirming data from a Danish study covering the years 2015-2019 (44). This is further in accordance with experimental evidence showing no vancomycin resistance transfer *via* membrane vesicles (45) and genomic analyses confirming lineage-specific *vanA*-plasmids (46). Although acquisition of *van* elements to generate novel VRE types might not be a very frequent event, our data showed that it happened occasionally. From an evolutionary perspective, such events, even if they are very rare, may lead to a fast clonal expansion of certain novel strain types that appear by chance, i.e., a “bottleneck effect.” In this context, extending genomic surveillance to include susceptible progenitor populations–such as VSE isolates from bloodstream infections, as in our case–has a clear and well-founded rationale. One example is ST117/CT2035, a van-negative lineage with consistent detection of multiple virulence gene variants typically found in hospital-adapted strains.

We identified outbreak clusters using cgMLST with ≤15 alleles threshold, which is routinely applied by the National Reference Centre for Enterococci for genomic surveillance and outbreak detection of *E. faecium* isolates (12, 47), revealing four dominant clusters (clusters 1-4). A commonly used ≤20 allele threshold (23) has been shown to lack sufficient discriminatory power for *E. faecium*, leading to the clustering of epidemiologically unrelated isolates and reduced resolution of transmission links (48). Recent studies advocate stricter clustering thresholds of ≤3 alleles to enhance discriminative power (49, 50); however, this conservative cut-off was derived for short-lived, spatially restricted outbreaks and may be too stringent for prolonged transmission events such as those analyzed here, where epidemiologically and phylogenetically linked cases naturally accumulate polymorphisms over time (51). We therefore applied an intermediate threshold of ≤15 alleles, which provides a balanced trade-off between sensitivity (detecting prolonged transmission events) and specificity (avoiding artificial clustering of unrelated lineages).

High-resolution genomic typing is increasingly recommended for hospital outbreak investigations (49). While cgMLST provides robust initial clustering, SKA enables refined discrimination, as demonstrated by Maechler *et al.* (48). We retrospectively applied SKA to ST117/CT71 isolates (cgMLST cluster 2), the main *vanB* peak driver. SKA resolved cgMLST cluster 2 into eight sub-clusters and 31 singletons, underscoring its high discriminatory power. This does not reflect an increase in outbreak-related isolates, but rather a higher-resolution subdivision of previously defined cgMLST clusters. Only one of eight SKA-defined clusters lacked likely epidemiological connections (either indicative of potential transmission or isolates originating from the same patient). The treating specialties of patients in cgMLST cluster 2 were consistent with previous studies identifying visceral surgery as a risk factor for nosocomial VRE infections (52), and internal medicine as having the highest incidence of VRE BSIs across six German university hospitals (33). Analysis of longitudinal patient-movement data identified epidemiological links involving 29 of 47 patients whose isolates had been classified as SKA singletons. These epidemiological connections, based on shared ward exposures, would have been difficult to detect manually due to repeated admissions and intra-hospital transfers over extended periods. Notably, isolates classified as SKA singletons exhibited significantly longer median intervals between linked BSI cases compared to isolates within SKA-defined clusters, suggesting that the 7-SNP SKA cutoff may be overly stringent for retrospective population studies involving prolonged transmission intervals. In contrast, the ≤15 allele cgMLST threshold may overestimate relatedness, potentially clustering genetically distinct isolates and generating false positives. Consequently, a tiered approach integrating high-resolution methods (such as SKA), cgMLST clustering, and detailed epidemiological analyses is necessary to balance sensitivity and specificity when investigating potential transmission events. While our findings align with the two-step genomic analysis pipeline proposed by Higgs *et al*. for investigating suspected VRE transmissions in real-time (49), our results highlight the importance of calibrating thresholds to the specific surveillance context.

Several limitations of this study should be acknowledged. Due to its retrospective design, comprehensive assessment of epidemiological links was not possible, particularly with regard to the intestinal colonization status of patients. As a result, we could not determine the number of nosocomially acquired isolates. Furthermore, detailed patient-movement data had to be extracted manually from the hospital information system; to keep the effort within realistic limits, we retrieved these data only for the largest cgMLST cluster (cluster 2), as it was most relevant to the *vanB* peak. Other infection sites such as urinary tract infections, together with colonization sources (e.g., fecal isolates) and environmental reservoirs, on which VRE can persist for extended periods (53), may have contributed to transmission but could not be systematically investigated in this retrospective study. Recent evidence indicates that environmental reservoirs can account for approximately one-third of potential VRE transmission events, with frequent detection on high-contact sites such as patient chairs and surrounding surfaces (54). Other studies have shown that VRE subtypes shared by multiple patients are often linked to contamination of communal bathrooms and medical devices, underscoring the role of the inanimate environment in sustaining transmission chains (55). Moreover, a recent comprehensive investigation from a Danish hospital has shown that nearly all epidemiological links for *E. faecium* within the hospital can be detected when isolates from all potential sources are analyzed (56). The study was conducted at a single tertiary care center, which may limit the generalizability of the findings to other healthcare settings.

In addition, isolate availability varied across study years, particularly in 2019 and 2021, potentially affecting the robustness of the population analysis during these periods.

Despite these limitations, our study demonstrates complex shifts in clonal lineages of *E. faecium* and *van* genotypes at Hannover Medical School. The *vanB* peak observed in 2019 most likely reflected a shift in the overall population structure of *E. faecium*. This shift was evident in the BSI isolates analyzed here, which displayed an oligoclonal pattern dominated by ST117/CT71, along with other *vanB-*positive clonal lineages, such as ST117/CT36 and ST117/CT1917. Of note, we identified clusters of *van*-negative lineages (ST117/CT929 and ST117/CT2505). Such clusters of *van*-negative *E. faecium* may be perceived as less concerning by clinicians due to available treatment options, but they have to be addressed by IPC measures similar to those applied for VRE. In genomic outbreak analysis, SKA and cgMLST provide complementary strengths, but their limitations require careful consideration, as their outcomes have direct implications for outbreak detection, reporting, and management. Adopting an integrated approach combining both methods will enhance genomic surveillance and provide valuable insights to guide targeted infection control strategies.

## Data availability

All raw sequencing reads generated in this study have been deposited in the European Nucleotide Archive (ENA) under the project accession number PRJEB91895. Raw data related to the investigation of epidemiological links and additional metadata for the whole-genome sequences cannot be shared publicly, as these data contain confidential patient information protected by the German Data Privacy Act, as well as by the ethics committee and the data protection commissioner of Hannover Medical School. Patient-related data, such as ward of admission, age, sex, underlying disease, or length of stay, are considered indirect identifiers and could enable re-identification of patients. To protect patient confidentiality and participants’ privacy, only anonymized and aggregated data can be made available. Researchers who meet the criteria for access to confidential data may request anonymized data by contacting the data protection commissioner of Hannover Medical School and the corresponding author. Data sharing is subject to approval by the ethics committee and the data protection commissioner.

## Acknowledgements

We thank the healthcare personnel on the wards for obtaining the patient samples, as well as the technical staff of the Institute for Medical Microbiology and Hospital Epidemiology for isolating the clinical strains and maintaining the strain collection. We thank Carola Fleige, Christine Bornstädt and the staff at MF2-Genome Sequencing and Genomic Epidemiology Core of the Robert Koch Institute for excellent technical assistance. We also thank PD Dr. Marius Vital for critically discussing the scope of the study.

This research received no specific grant from any funding agency in the public, commercial, or not-for-profit sectors.

