## Supplementary material for "Genomic Epidemiology of *Enterococcus faecium* Bloodstream Infections During a VanB-type VRE Peak Reveals an Oligoclonal Scenario: An Observational Study at a German University Hospital (2017–2022)": Figure S1 and Figure S2

**SUPPLEMENTARY FIGURE 1. Heatmap of virulence factors in the most prevalent complex types (CTs).** Columns represent virulence genes and variant calls as defined by *VirulenceFinder* (Center for Genomic Epidemiology, Technical University of Denmark). Each row corresponds to one of the most prevalent sequence type (ST)-CT combinations identified in this study. Color intensity indicates the percentage (0–100%) of isolates carrying each variant, with darker shades representing higher percentages. If an isolate matched more than one variant in the database with identical identity and gene coverage, this was defined as an *ambiguous call*. In the heatmap, only the percentage for the dominant variant within each CT is shown. Abbreviations: Efm, *E. faecium*; Elts, *E. lactis*; PGC, pili gene cluster; PTS, phosphotransferase system.

**SUPPLEMENTARY FIGURE 2. Timeline of the VRE BSI cases assigned to cgMLST cluster 2.** Each row represents a single patient (n=47), grouped by the treating specialty. These specialties were categorized into three organ system–related groups (see Materials and Methods). The timeline spans the whole study period (monthly ticks). Patients with a blue-colored patient identifier had a known intestinal VRE colonization prior to or at the time of BSI. The SKA-Cluster column lists the SKA clusters (11-18) assigned to each patient's isolate(s). Colored boxes (or narrow vertical bars for short stays) indicate the hospital ward(s) where each patient was admitted during the corresponding time period; each color denotes a distinct ward. The letter "i" marks the date the VRE-positive blood culture was obtained.

**Supplementary Figure 1**

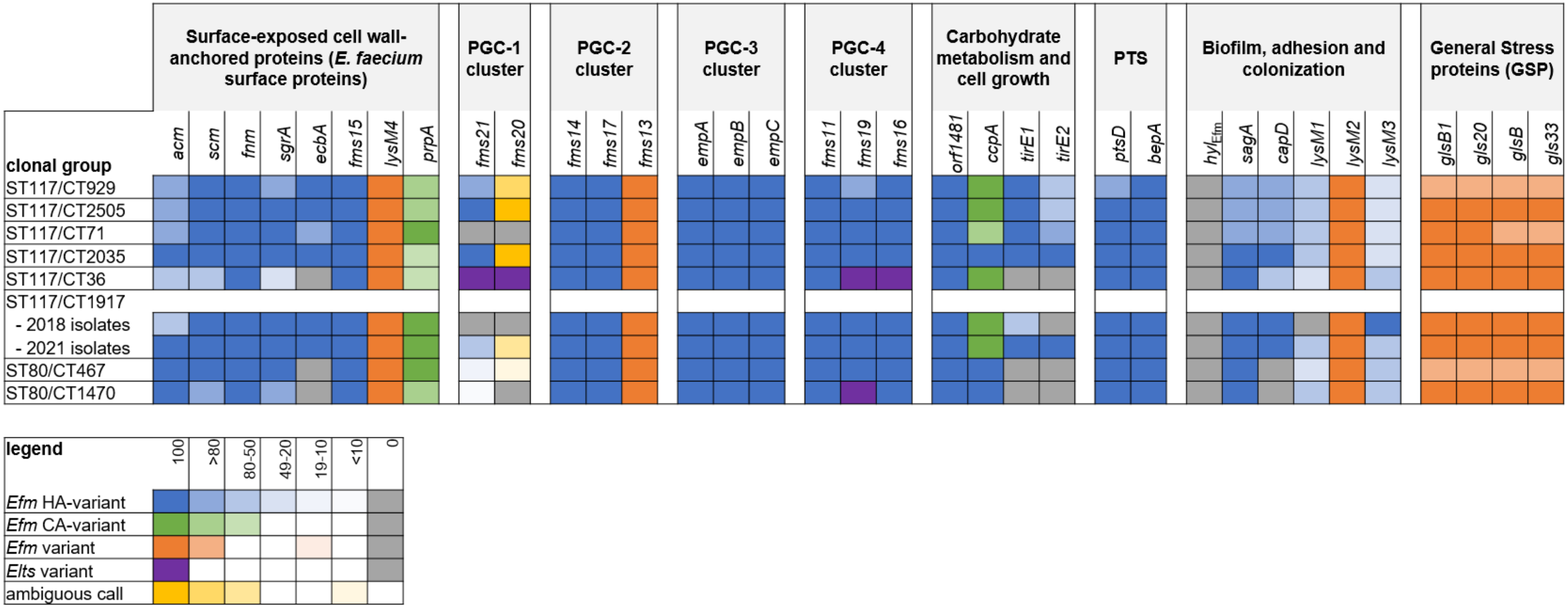

### Supplementary Figure 2

| Underlying disease (organ system) | Patient pseudonym | SKA-Cluster |
|---|---|---|
| Excretory system | 12346 | |
| Excretory system | 11702 | |
| Excretory system | 10985 | 14 |
| Excretory system | 10158 | |
| Excretory system | 10860 | |
| Excretory system | 11793 | |
| Excretory system | 11668 | 11 |
| Excretory system | 13067 | |
| Excretory system | 13269 | 12 |
| Excretory system | 10113 | |
| Excretory system | 11675 | 12 |
| Excretory system | 13490 | |
| Excretory system | 12325 | |
| Excretory system | 11149 | 11 |
| Excretory system | 12304 | 13 |
| Excretory system | 10716 | 13 |
| Excretory system | 12636 | 17 |
| Excretory system | 10466 | |
| Excretory system | 11894 | |
| Excretory system | 12650 | |
| Hematopoietic system | 11171 | |
| Hematopoietic system | 12037 | 14 |
| Hematopoietic system | 10701 | 14 |
| Hematopoietic system | 12982 | 15 |
| Hematopoietic system | 11810 | 15 |
| Hematopoietic system | 10278 | |
| Hematopoietic system | 12711 | |
| Hematopoietic system | 10271 | 16 |
| Hematopoietic system | 12641 | 16 |
| Hematopoietic system | 11327 | |
| Hematopoietic system | 12907 | 18 |
| Hematopoietic system | 12331 | |
| Hematopoietic system | 12712 | |
| Hematopoietic system | 12256 | |
| Other | 10566 | |
| Respiratory and cardiovascular system | 12875 | 13 |
| Respiratory and cardiovascular system | 13122 | |
| Respiratory and cardiovascular system | 12229 | |
| Respiratory and cardiovascular system | 11304 | 12 |
| Respiratory and cardiovascular system | 13007 | |
| Respiratory and cardiovascular system | 11002 | |
| Respiratory and cardiovascular system | 11089 | 11 |
| Respiratory and cardiovascular system | 12854 | |
| Respiratory and cardiovascular system | 11311 | |
| Respiratory and cardiovascular system | 12457 | |
| Respiratory and cardiovascular system | 11830 | |
| Respiratory and cardiovascular system | 12916 | |

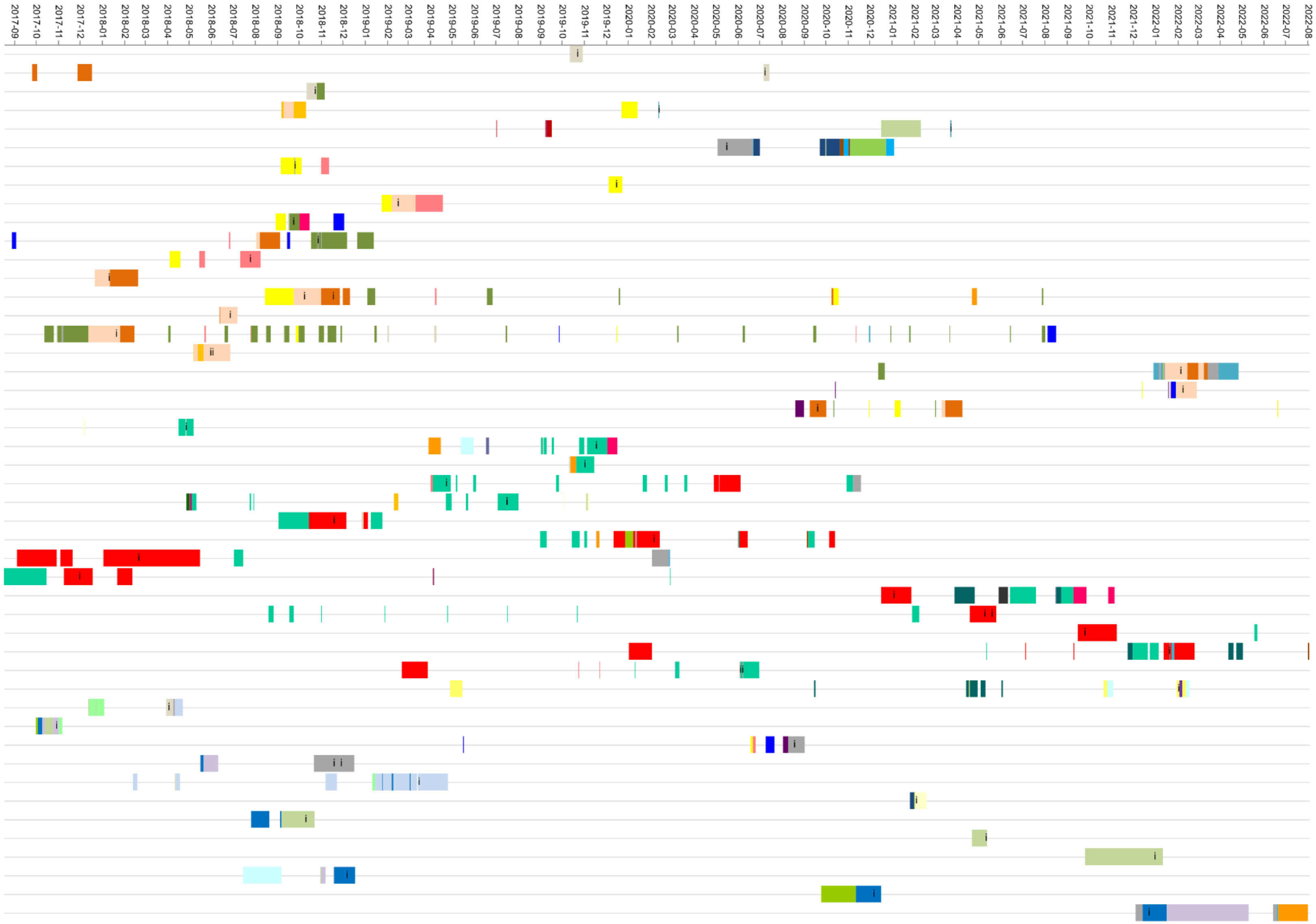
